# Convergence of Cancer Mortality Rates Across U.S. States: The Role of Socioeconomic and Behavioral Risk Factors

**DOI:** 10.1101/2025.05.12.25327403

**Authors:** Alberto Carlo Olivei

## Abstract

**Background:** Geographic disparities in U.S. cancer mortality rates raise concerns about whether different states are equally benefiting from advances in best practices for cancer prevention, early detection, and care. This study assesses whether U.S. states are catching up to the frontier of best practices while accounting for state-level differences in cancer risk factors.

**Methods:** We analyze age-adjusted cancer mortality rates across 48 U.S. states and evaluate their convergence over the period from 1997 to 2021. Specifically, we examine whether states with initially higher mortality rates have experienced steeper declines in those rates over time, both with and without adjusting for state-level differences in cancer-related risk factors. We also investigate whether risk factors themselves have converged over time.

**Results:** There is little evidence of unconditional convergence in cancer mortality across U.S. states. However, when controlling for risk factors such as smoking, obesity, share of manufacturing employment, and per capita GDP, convergence becomes evident. Conditional on those risk factors being similar, if one state’s mortality rate is higher than another’s, then roughly half of the gap closes within 12 years. Persistent disparities in cancer-related risk factors largely explain the absence of unconditional convergence.

**Conclusions:** These findings suggest states are making progress in best practices for cancer care. However, to reduce the dispersion of state-level cancer mortality rates around the national average, it will be necessary to address enduring state-level disparities in socioeconomic and behavioral determinants of cancer mortality.

## 1. Introduction

Cancer mortality rates in the United States have declined since the 1990s. [1] However, demographic and geographic disparities persist, presenting a significant health challenge. [2-4] As best practices in cancer prevention, early detection, and treatment continue to diffuse nationwide, one might expect these disparities to narrow. [5] This study evaluates whether cancer mortality rates across U.S. states have converged between 1997 and 2021, consistent with progress toward the widespread adoption of effective cancer control strategies. We also analyze factors that contribute to persistent differences in cancer mortality rates across states, despite the broad dissemination of knowledge and care. A better understanding of the factors that enhance or impede convergence can help guide policy efforts aimed at reducing disparities in cancer outcomes across the country. [6,7]

## 2. Data and Sample

This study examines cancer mortality rates across 48 contiguous U.S. states (excluding D.C.) over six four-year periods, starting in 1997. The periods are 1997-2001, 2001-2005, …, up to 2017-2021. The sample includes 288 observations (48 states times 6 periods). The start date is constrained by state-level GDP data availability. Age-adjusted state-level cancer death rates are obtained from the CDC Wonder database (ICD-10 codes C00-C97 from 1999 onward, and ICD-9 codes 140-208 for 1997-1998). State-level population data are sourced from the same dataset. State-level data on the percentages of obese individuals and everyday smokers come from the CDC Behavioral Risk Factor Data Portal. Real, or inflation-adjusted, state-level GDP data have been sourced from the Bureau of Economic Analysis and transformed on a per capita basis. State-level manufacturing employment data, expressed as a percentage of total employment, come from the Bureau of Labor Statistics Current Employment Statistics. The discussion section also mentions results not reported in the text that utilize additional controls, which consist of state-level Black population, the percentage of college-educated individuals (both from the CDC Wonder database), and the poverty rate (from the Bureau of the Census). All data used in this study are de-identified and publicly available.

## 3. Measures

The primary outcome is the four-year change in the log of age-adjusted cancer mortality rates at the state level, calculated as the log death rate at the end of each period minus the log death rate at the beginning. For example, for the 1997–2001 period, the outcome is the log mortality rate in 2001 minus that in 1997. The change is explained using the initial mortality rate at the start of the period (e.g., 1997 for the 1997–2001 period), along with a set of state-level controls also measured at the beginning of each period. These include the log-transformed values of the percentage of everyday smokers, the percentage of the population classified as obese, the share of employment in manufacturing, and per capita real GDP. This approach is applied across all six periods.

## 4. Statistical analysis

The analysis seeks to determine whether states with higher initial cancer mortality rates catch up to those with lower rates. Our approach is common in the demographic literature examining convergence in mortality rates across regions over time. [8,9] We conduct ordinary least squares (OLS) regressions using pooled data from 48 states over six non-overlapping four-year periods. The dependent variable is the four-year change in the log of the cancer mortality rate; explanatory variables include the log of the initial mortality rate and a set of log-transformed state-level controls measured at the start of each period. These regressions include constants for each time period, which account for changes in averages across states over time.

To assess convergence in the control variables themselves, we run separate OLS regressions for each control, using the four-year change as the dependent variable and the initial value as the explanatory variable, again with period indicators. The statistical analysis uses a two-sided p-value of .05. All regressions were executed using the reg command in Stata 18.

## 5. Results

Figure 1 presents a boxplot of log cancer mortality across states from 1997 to 2021. Cancer mortality rates declined nationally – the median state-level rate fell by approximately 32%. However, the dispersion across states remained stable. The standard deviation of the log cancer mortality rate was 0.0870 in 1997 and 0.0967 in 2021, indicating no clear trend toward convergence. The first column of Table 1 confirms this pattern by regressing the 4-year change in state-level cancer mortality on the initial state-level mortality for the six periods from 1997 to 2021, while also controlling for period-specific effects (not reported in the table). A reduction in dispersion would be consistent with a negative coefficient for initial rates, implying that states with higher mortality would experience greater declines. The estimated coefficient on the initial rate is −0.035 (SE = 0.022, p = 0.117), suggesting only weak and statistically insignificant convergence as the gap in initial mortality closes by roughly 15% over two decades – an extremely slow pace. This result provides little evidence of unconditional convergence.

**Table 1.** Estimates of Conditional and Unconditional Convergence in Cancer Mortality Rates. The dependent variable is the 4-year change in the state-level log cancer mortality rate for the periods 1997-2001, 2001-2005, 2005-2009, 2009-2013, 2013-2017, and 2017-2021.

|  | (1) Without Cancer Risk Factors | (2) With Cancer Risk Factors |
| --- | --- | --- |
| Initial Cancer Mortality Rate | -0.0355<br>SE = 0.0222, \|p\| = 0.117 | **-0.2237**<br>SE = 0.0350, \|p\| < 0.001 |
| Initial Tobacco Use |  | **0.0735**<br>SE = 0.0167, \|p\| < 0.001 |
| Initial Obesity |  | **0.0541**<br>SE = 0.0154, \|p\| = 0.001 |
| Initial Share of Manufacturing |  | **0.0113**<br>SE = 0.0029, \|p\| < 0.001 |
| Initial Per-Capita GDP |  | **-0.0245**<br>SE = 0.0079, \|p\| = 0.003 |
| Number of Observations | 288 | 288 |
| R-squared | 0.217 | 0.380 |

Note: SE = standard errors. Standard errors are clustered at the state level. Indicator variables (not reported) for each of the six periods considered are included in the analysis. Estimates in bold indicate that the 95 percent confidence interval does not include zero. All explanatory variables are expressed in logs. For example, in column (2), the value of -0.2237 means that a one percentage point higher initial cancer mortality rate lowers the following 4-year change in mortality rate by 0.2237 of one percentage point.

**Figure 1.**
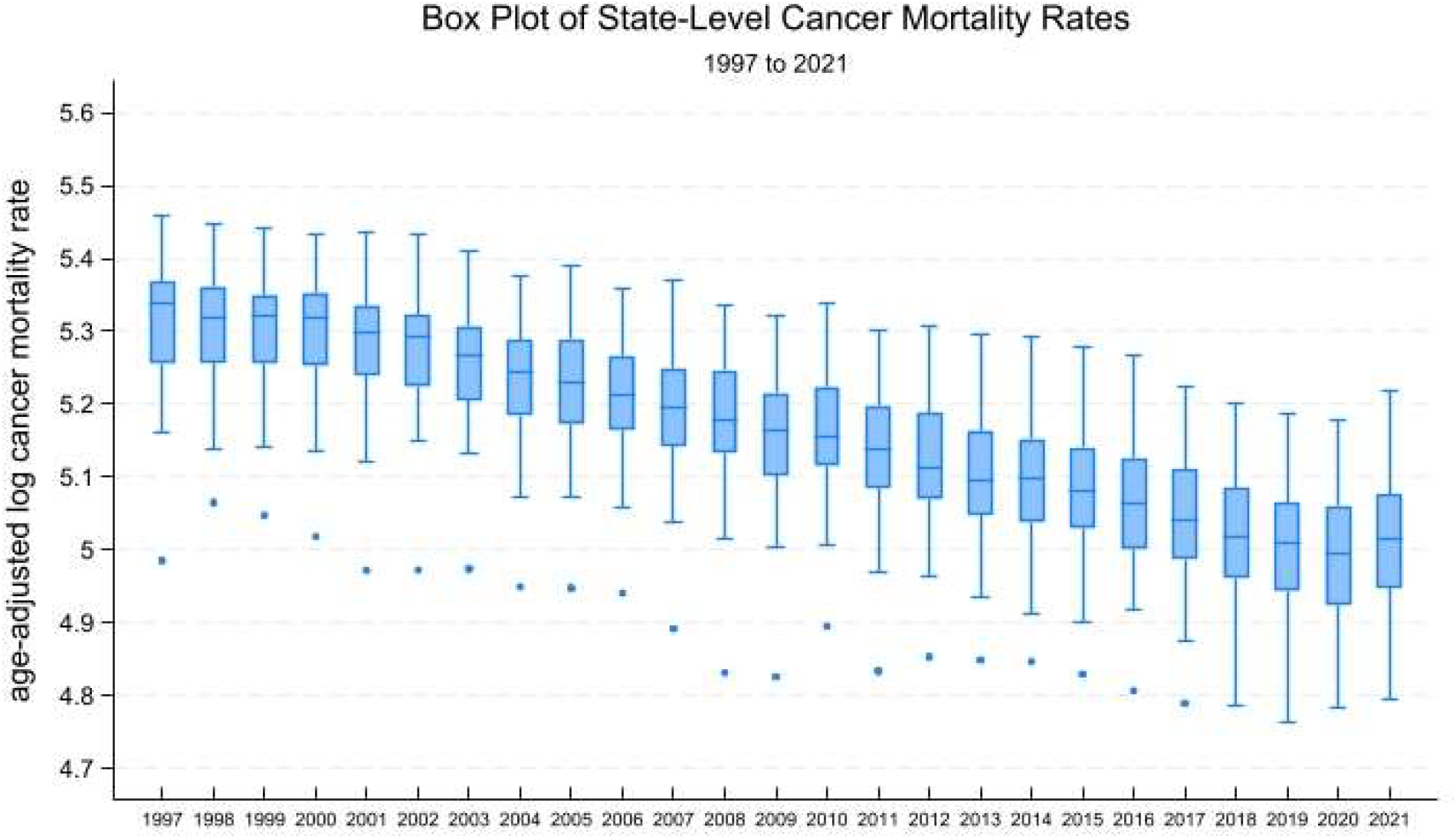
Box plot distribution of age-adjusted log cancer death rates for the 48 contiguous US states (excluding DC) from 1997 to 2021. Each box displays the median log cancer mortality rate for a specific year (represented by the line in the box), while the upper and lower ends of the box indicate the 75th and 25th percentiles, respectively. The whiskers represent the upper and lower adjacent values, defined as the third quartile plus 1.5 times the interquartile range (upper adjacent value) and the first quartile minus 1.5 times the interquartile range (lower adjacent value). Dots represent values outside of the range spanned by the box and whiskers.

The next column shows how convergence findings change when controlling for factors affecting cancer mortality disparities across states, such as tobacco usage, obesity, the share of manufacturing workers, and per capita GDP. Tobacco and obesity are established cancer risk factors. [10,11] High occupational exposure to carcinogens in manufacturing and proximity to these activities may also increase cancer risk. [12] The connection between income and cancer death rates could relate to less affordable and less available care. [13] Estimates for these controls have expected signs: tobacco usage, obesity, and manufacturing share correlate positively with changes in cancer mortality, while real per capita GDP correlates negatively. Notably, the coefficient on initial mortality becomes substantially larger in magnitude (−0.223, SE = 0.035, p < 0.001), implying a much stronger convergence pattern. Conditional on these risk factors, nearly half of the mortality gap between two otherwise similar states closes within approximately 12 years. This suggests that the diffusion of effective cancer control practices is contributing to convergence once differences in structural and behavioral risk factors are taken into account.

Table 2 illustrates one reason for the difference in cancer death rates’ unconditional and conditional convergence across states. It presents estimates from regressing the four-year change on the initial value for each control, accounting for period-specific effects. Tobacco usage, the share of manufacturing, and real GDP have not converged over time. There is evidence of convergence in obesity trends; however, these trends have been on the rise. [14] In short, the fact that some important cancer risk factors have not equalized across states accounts for the persistence of cancer mortality dispersion. Still, conditional on considering these factors, state-level differences in cancer mortality rates have been narrowing.

**Table 2.**
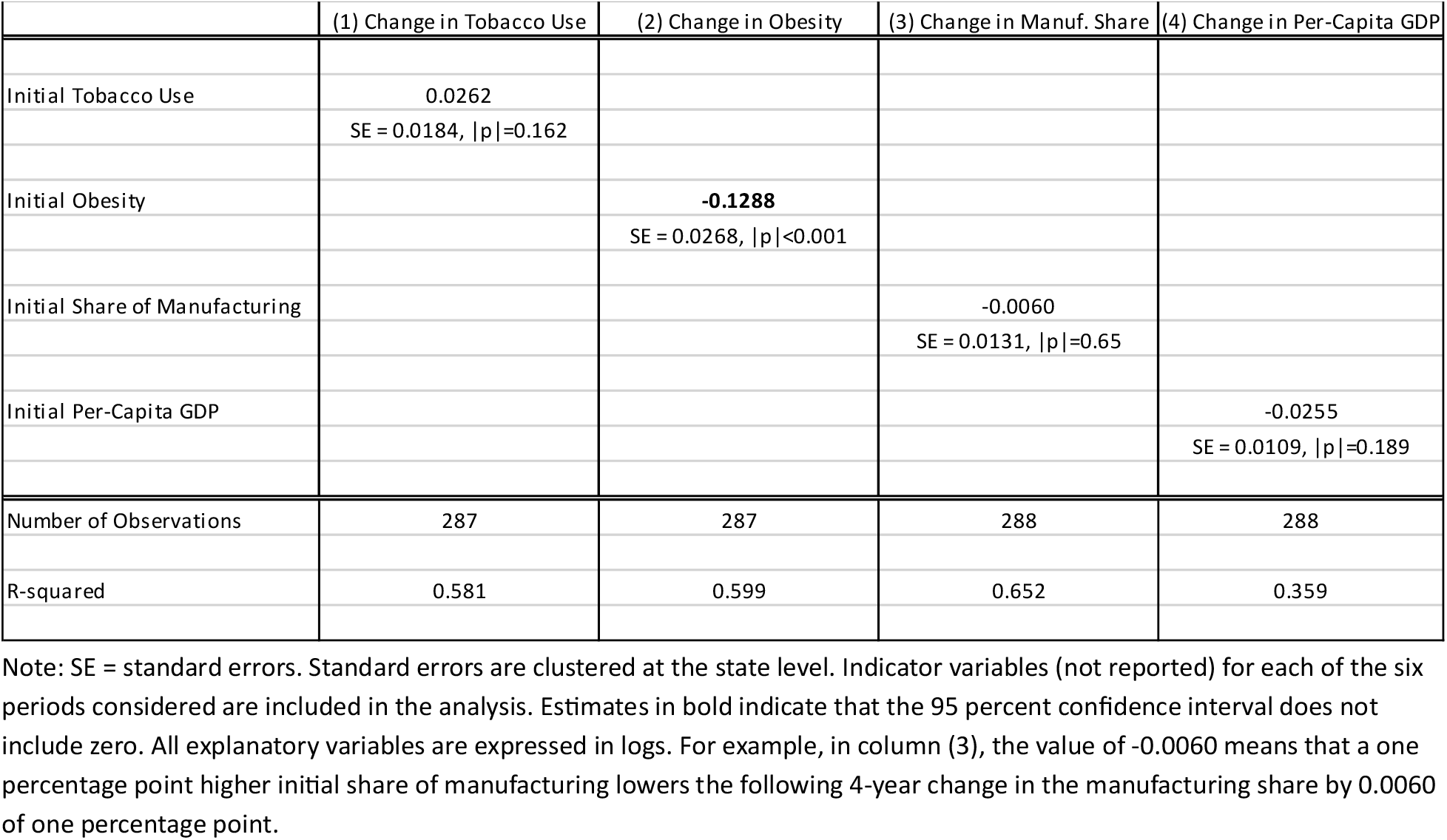
Estimates of Unconditional Convergence in Cancer Risk Factors. The dependent variable is the 4-year change in each specific cancer socio-economic risk factor, for the periods 1997-2001, 2001-2005, 2005-2009, 2009-2013, 2013-2017, and 2017-2021.

## 6. Discussion

Cancer remains the second leading cause of death in the United States, and while national mortality rates have declined since the 1990s, geographic disparities persist. [1] This study explores the persistent dispersion in cancer mortality rates across U.S. states. We analyze data from 1997 to 2021 for the 48 contiguous states (excluding D.C.) at four-year intervals. This interval length is suitable for assessing convergence in cancer mortality rates, and the repeated cross-section across six distinct four-year periods provides enough observations to control for specific state factors that influence cancer mortality rates.

Three key findings emerge from the analysis. First, from 1997 to 2021, there was a lack of unconditional convergence in cancer mortality rates among states. Second, when accounting for differences in smoking prevalence, obesity, manufacturing employment, and per-capita GDP, a strong pattern of conditional convergence emerges. States with similar levels of these risk factors tend to converge toward comparable mortality rates, suggesting that advancements in cancer treatment are being adopted nationally and benefiting states with historically higher mortality. Third, the persistence of geographic disparities is largely driven by the lack of convergence in these underlying risk factors themselves, reinforcing a pattern of divergence in health outcomes where these conditions remain uneven.

One important limitation of this study is that other factors influencing cancer mortality may vary across states and were not considered. While the analysis could expand to include additional covariates, including the state-level share of the Black population [15], college-educated individuals, and poverty rate, did not affect the findings. At least in this setting, these additional variables do not yield additional explanatory power (see Appendix), perhaps because they tend to correlate with GDP. Nonetheless, other omitted variables could still impact the rate of conditional convergence in cancer mortality.

## 7. Conclusion

Geographic disparities in cancer mortality across the U.S. persist, raising the important question of whether all states are effectively leveraging advancements in best practices for cancer prevention, early detection, and care. [7] This study finds that states with similar levels of tobacco use, obesity, manufacturing employment, and per-capita GDP tend to converge to the same cancer mortality rate – suggesting that progress is reaching lagging states. However, broader convergence remains hindered by persistent disparities in these underlying risk factors, many of which show little evidence of narrowing. Efforts to reduce cancer mortality disparities must therefore prioritize targeted strategies to close gaps in the underlying social and behavioral drivers of cancer risk across states.

## Data Availability

Data and replications files are available at: https://github.com/ACOlivei/US-States-Cancer-Mortality.

## Financial disclosure

No funding was received for this study.

## Declaration of competing interest

The author reports no conflict of interest.

## Appendix

**Table 3.** Estimates of Conditional Convergence in Cancer Mortality Rates with Additional Controls. The dependent variable is the 4-year change in the state-level log cancer mortality rate for the periods 1997-2001, 2001-2005, 2005-2009, 2009-2013, 2013-2017, and 2017-2021.

|  | (1) Baseline | (2) With Additional Controls |
| --- | --- | --- |
| Initial Cancer Mortality Rate | **-0.2237**<br>SE = 0.0350, \|p\| < 0.001 | **-0.2089**<br>SE = 0.0357, \|p\| < 0.001 |
| Initial Tobacco Use | **0.0735**<br>SE = 0.0167, \|p\| < 0.001 | **0.0676**<br>SE = 0.0172, \|p\| < 0.001 |
| Initial Obesity | **0.0541**<br>SE = 0.0154, \|p\| = 0.001 | **0.0504**<br>SE = 0.0191, \|p\| = 0.011 |
| Initial Share of Manufacturing | **0.0113**<br>SE = 0.0029, \|p\| < 0.001 | **0.0109**<br>SE = 0.0025, \|p\| < 0.001 |
| Initial Per-Capita GDP | **-0.0245**<br>SE = 0.0079, \|p\| = 0.003 | -0.0197<br>SE = 0.0123, \|p\| = 0.116 |
| Initial Black Population Share |  | -0.0013<br>SE = 0.0014, \|p\| = 0.421 |
| Initial College Share |  | -0.0174<br>SE = 0.0159, \|p\| = 0.282 |
| Initial Poverty Rate |  | -0.0085<br>SE = 0.0108, \|p\| = 0.435 |
| Number of Observations | 288 | 288 |
| R-squared | 0.380 | 0.385 |

Note: SE = standard errors. Standard errors are clustered at the state level. Indicator variables (not reported) for each of the six periods considered are included in the analysis. Estimates in bold indicate that the 95 percent confidence interval does not include zero. Column (1) in the table replicates results reported in Table 1. Column (2) adds additional socio-economic cancer risk factors. All explanatory variables are expressed in logs. For example, in column (1), the value of -0.2237 means that a one percentage point higher initial cancer mortality rate lowers the following 4-year change in mortality rate by 0.2237 of one percentage point.

## Notes

### Competing Interest Statement

The authors have declared no competing interest.

### Funding Statement

This study did not receive any funding

### Summary of Updates

Revision to correct a misspelling in author's affiliation in the cover page.

